# Design-Induced Circularity in Alzheimer’s Biomarker Trials and Trial-Ready Cohorts: Enrichment, Data-Driven Selection, and Estimand Mismatch

**DOI:** 10.64898/2025.12.20.25342753

**Authors:** Khushboo Verma, Satwant Kumar

## Abstract

**Background:** Biomarkers increasingly gate Alzheimer disease trials and clinical access, raising risk of design-induced bias when biomarker data are reused for both selection and inference.

**Objective:** To quantify circularity patterns in Alzheimer disease biomarker trials and trial-ready cohorts and, using mechanistic experiments, estimate effects on discrimination, calibration/transportability, and false-positive risk.

**Methods:** We audited PubMed-indexed biomarker interventional trials and trial-ready cohorts (2020-2024) using dual independent full-text review and mechanism classification. Two simulation experiments examined (1) enrichment-induced range restriction with transport to an unselected population and (2) selection-inference reuse (“double dipping”) versus split-sample confirmation.

**Results:** Among 72 studies, 10 (13.9%) raised possible/definite non-independence; enrichment/range restriction predominated (7/10). In simulation experiment 1, enrichment preserved nominal type I error under the null but shifted the estimand under signal, attenuating discrimination and degrading calibration when enriched-fit models were applied to unselected populations. In simulation experiment 2, double dipping produced *P*(*p* < 0.05) = 0.994 under the null (*n* = 200, *p* = 100) versus 0.047 with split-sample confirmation, with winner’s-curse effect inflation.

**Conclusions:** Circularity was uncommon but nontrivial and most often reflected enrichment with implicit generalization beyond the enriched population. Safeguards include explicit target populations/estimands and separation of discovery from confirmation (or prespecified multiplicity control), with clear validation and leakage prevention when biomarker claims are intended to guide decisions.

## Introduction

Disease-modifying and prevention trials in Alzheimer’s disease (AD) increasingly embed biomarker confirmation and staging into enrollment and outcomes, using CSF and plasma amyloid-β (Aβ) and phosphorylated tau, amyloid and tau PET, and trial-ready recruitment infrastructures to enrich for AD biology and to quantify pharmacodynamic and prognostic signals [1–7]. As scalable blood-based biomarkers mature, they are also being positioned to support prescreening and risk stratification for trial recruitment and early-stage disease detection [8–10]. Consequently, biomarker choices increasingly determine who is studied and condition how “biological response” is interpreted in regulatory and clinical implementation debates, including discussions about surrogate endpoints and treatment eligibility pathways [5, 6, 11, 12].

These advances create genuine opportunities for efficient trial conduct, but they also introduce design-induced non-independence: distortions driven by conditioning on biomarker status (eg, eligibility, enrichment, or thresholded subgrouping) and by downstream analytic choices, rather than by random sampling variability alone [13, 14]. In particular, two mechanisms are most relevant. Enrichment-related non-independence arises when biomarker criteria restrict the analyzed sample and subsequent associations or performance estimates are implicitly discussed as if they apply to the broader clinic-source population [13, 14]. Selection-related non-independence (“double dipping”) arises when many candidate biomarkers, regions, or cut points are screened in a dataset and the same dataset is then used for inference on the selected “winner,” producing optimistically biased p values, effect sizes, and performance estimates unless selection and evaluation are properly separated or nested within validation [15–19].

One recurrent problem is enrichment-related circularity: eligibility is restricted using a biomarker, but conclusions are later framed as if they apply to a broader clinical population. Many trials and trial-ready cohorts require amyloid positivity on PET, abnormal plasma/CSF profiles, or elevated composite risk scores for entry, then report biomarker-outcome associations or prognostic performance as though they generalize to all patients with cognitive complaints. In these designs, p-values and internal discrimination metrics can look acceptable, yet the target estimand has shifted, from an unselected clinic population to a biomarker-defined subset, without being stated explicitly [11, 14]. Because enrichment changes the case mix, it can alter risk gradients, discrimination, and calibration when models are transported back to routine care, even when the underlying biology is unchanged [20, 21]. This concern aligns with broader methodological guidance emphasizing explicit estimand definition in clinical trials [22].

A second pattern is selection-related circularity (“double dipping”): many candidate biomarkers, regions of interest, or composite scores are screened within a single dataset, the strongest association is selected, and the corresponding p-value or effect size is then reported as though it arose from a single pre-specified test [16]. This form of circular analysis is well described in neuroimaging and other high-dimensional settings, where re-using the same data for selection and inference produces biased statistics, inflates false positives, and yields over-optimistic effect estimates [16]. Even when a true signal exists, selecting the most extreme result among many candidates induces winner’s-curse inflation, so reported effects tend to be exaggerated and less reproducible [23, 24]. As Alzheimer’s trials increasingly incorporate multi-marker panels, omics, and rich imaging, this selection-inference reuse becomes tempting and sometimes operationally convenient unless independence (e.g., split-sample discovery/confirmation) or appropriate multiplicity control is built into the design and analysis [25].

Despite these concerns, it remains unclear how often enrichment-related circularity, selection-related circularity, or hybrids occur in recent Alzheimer’s biomarker trials and trial-ready cohorts, because this typically requires manual, full-text assessment of eligibility criteria, analysis pipelines, and validation claims. Likewise, Alzheimer’s-specific quantitative bounds on the resulting distortions, changes in discrimination and calibration under enrichment, and false-positive/effect-size inflation under selection are not routinely reported under realistic design conditions [14]. This matters because biomarker claims are increasingly used to justify trial designs and endpoints, shape regulatory judgments (including biomarker-based surrogate reasoning), and influence payer and health-system decisions about whom to test and treat [22, 26].

In this study, we address design-induced bias in Alzheimer’s biomarker trials by combining manual full-text audit with mechanistic simulations. We first conduct a structured review of biomarker-enriched AD trials and trial-ready cohorts published between 2020 and 2024, classifying each study based on eligibility criteria, analytic workflow, and validation claims as showing definite circularity, possible circularity, or apparently independent inference. For studies with definite or possible circularity, we further identify the dominant pattern (enrichment-related, selection-related, or hybrid) and record co-occurring design features relevant to bias risk, including validation strategy, handling of multiplicity, and safeguards against information leakage [26, 27]. This approach allows us to move beyond abstract methodological concerns and to empirically characterize how often distinct forms of circular inference appear in contemporary Alzheimer’s biomarker research [22, 28, 29].

We then use two aligned simulation experiments to quantify how these patterns can distort inference under controlled, transparent assumptions. Experiment 1 represents a setting where a latent disease process influences both a clinical outcome and a biomarker; we compare (i) a “global” design sampling from the full population to (ii) a biomarker-enriched design that conditions analysis on individuals in the upper tail of the biomarker distribution. Because discrimination and calibration depend on case mix and the range of risk in the analyzed sample, enrichment can change apparent model performance and may degrade calibration when models are transported back to an unselected clinical population [20, 21, 30]. Experiment 2 represents a high-dimensional selection setting: many candidate biomarkers are screened, a “best” candidate is chosen, and inference is performed either in the same dataset (a double-dip pipeline) or in an independent confirmation set (a split-sample pipeline). Re-using the same data for selection and inference is expected to inflate false positives and overstate effects unless independence or multiplicity control is built into the workflow [16].

Taken together, our audit and simulation experiments highlight two complementary failure modes of circular inference in Alzheimer’s biomarker research. First, biomarker-based enrichment can create an implicit estimand mismatch, where associations or prognostic performance estimated in a biomarker-defined subset are later interpreted as if they apply to an unselected clinical population, despite well-known case-mix effects on discrimination and calibration [14, 20, 21]. Second, selection-inference reuse (double dipping) can generate apparently “significant” findings and exaggerated effects when many candidates are screened and the same dataset is used for both selection and testing [16]. Because biomarker claims increasingly inform trial design, endpoint reasoning, and downstream access decisions, these risks are not merely technical; they can influence what evidence is produced and how it is applied [31]. We therefore aim to provide practical guidance for biomarker-enriched AD studies, emphasizing (i) explicit statement of the target estimand and intended target population, (ii) separation of discovery from confirmation (or principled multiplicity control) when selection is unavoidable, and (iii) transparent reporting and bias-aware validation consistent with established prediction-model standards [14, 22, 26].

## Methods

### Identification of eligible studies

We performed a targeted methodological audit of recent Alzheimer’s disease (AD) biomarker trials and trial-ready cohorts using PubMed (MEDLINE) as the bibliographic source. Our aim was not a systematic review of treatment efficacy, but a focused assessment of how biomarkers are incorporated into study design and analysis in contemporary AD biomarker-enriched research. We implemented two complementary PubMed searches to capture (i) biomarker-enriched interventional trials and (ii) trial-ready observational infrastructures (registries, screening platforms, and cohorts explicitly designed to support trial recruitment). Searches were executed programmatically using the NCBI Entrez E-utilities interface and restricted to publications dated 2020-2024. Each search combined (a) AD/cognitive impairment terms, (b) fluid and imaging biomarker terms (plasma/CSF Aβ and tau; amyloid/tau PET; MRI; multimarker panels/risk scores), and depending on the search (c) trial design indicators (e.g., randomized/phase terminology) or trial-readiness terms (e.g., “trial-ready,” “screening,” “registry,” “recruitment cohort”). To focus on primary research, we excluded non-original publication types using PubMed publication-type filters and removed animal-only records using the standard PubMed MeSH species exclusion filter. Full Boolean search strings are available in the online repository (see subsection Data availability).

Records retrieved by either search were merged and deduplicated by PubMed identifier (PMID). We included full-length, peer-reviewed studies in humans that enrolled participants with AD, prodromal AD, or AD-related cognitive impairment and that were either (i) interventional trials (any phase, including prevention/platform designs) or (ii) trial-ready observational infrastructures (cohorts/registries/screening platforms explicitly intended to support AD trial recruitment). Eligible studies had to incorporate fluid and/or imaging biomarkers (e.g., plasma/CSF Aβ or tau; amyloid/tau PET; MRI; multimarker panels or composite scores) as part of eligibility/enrichment, as endpoints/key outcomes, and/or within prognostic or risk-modeling analyses. We excluded animal or in-vitro studies, purely basic imaging/omics work without an explicit trial/trial-readiness context, and non-original or non-full-text formats (e.g., reviews, editorials/letters, and conference abstracts without a peer-reviewed article). Two reviewers independently screened titles and abstracts for eligibility, followed by independent full-text assessment of all potentially eligible reports. Disagreements at either stage were resolved by discussion and, if needed, adjudication by a third reviewer. Study identification and selection are summarized in a PRISMA-style flow diagram (**Figure 1**).

**Figure 1:**
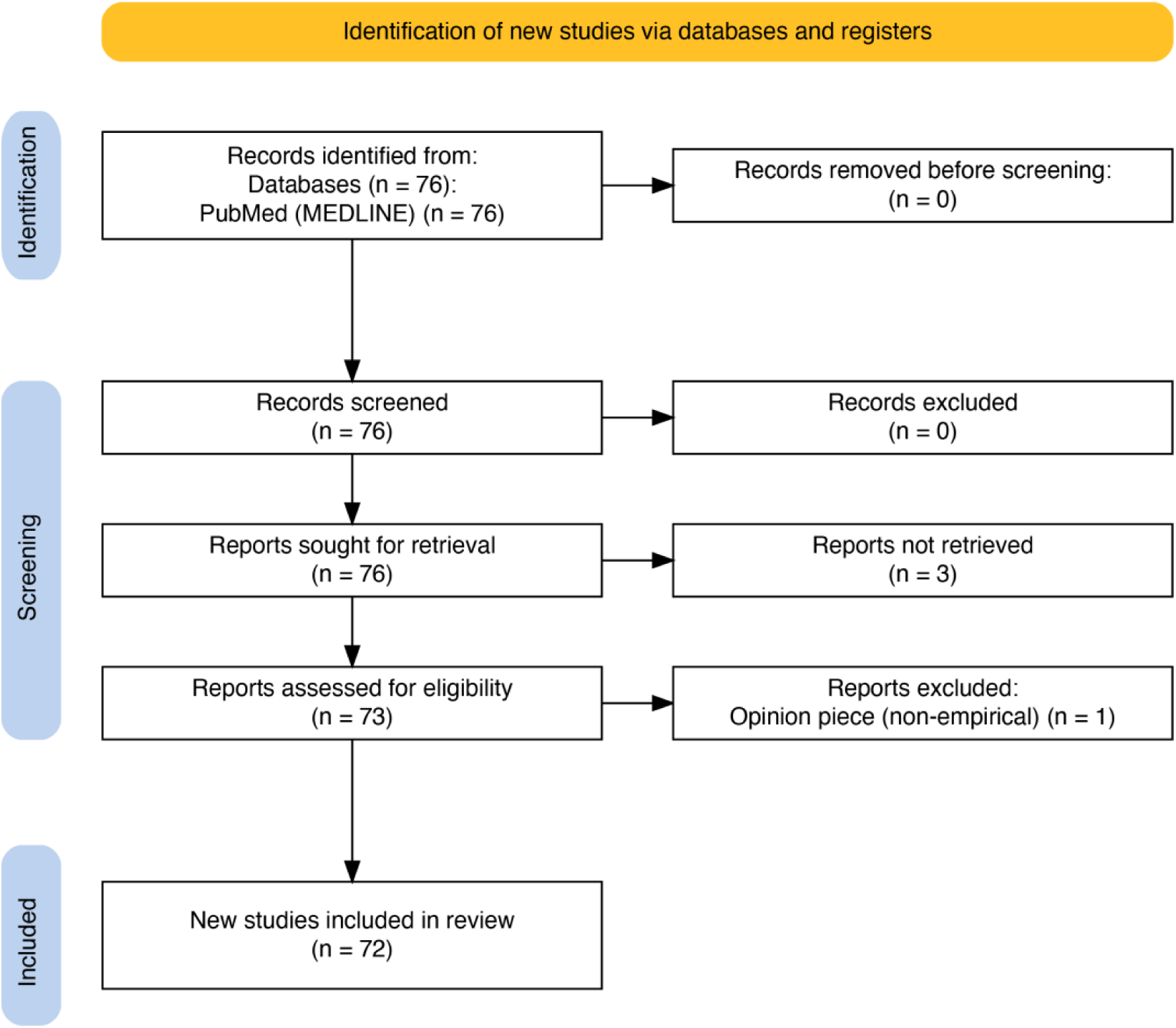
PRISMA flow diagram of study selection for the methodological audit. PubMed (MEDLINE) records (2020-2024) were identified using prespecified searches for Alzheimer’s biomarker trials and trial-ready cohorts. Seventy-six records were screened; 76 reports were sought for retrieval, of which 3 were not retrieved. Seventy-three full texts were assessed for eligibility; 1 non-empirical opinion piece was excluded. Seventy-two studies were included in the final audit.

Using a prespecified extraction template, two reviewers (authors) independently abstracted key study characteristics (design and setting, sample size and eligibility criteria, biomarker modalities, biomarker roles in eligibility/endpoints/prediction, and primary analytic approach). All coding and judgments were based on manual full-text review. Discrepancies were resolved by consensus.

We defined circularity as any design or analysis pattern in which biomarker information is used to shape the analyzed sample or select features, and the same biomarker signal is then used to support claims about predictive, surrogate, or diagnostic performance without appropriate separation, independent confirmation, or explicit accounting for selection. Each study was classified as definite circularity, possible circularity (insufficient reporting to decide), or apparently independent. For studies with definite/possible circularity, we further assigned the dominant pattern: enrichment-related, selection-related (double dipping), or hybrid. Two reviewers coded all studies independently; disagreements were resolved by consensus. Inter-rater agreement for the binary primary circular vs non-circular classification was summarized using Cohen’s κ (0.76).

To characterize factors that directly modulate circularity-related bias, we coded two additional co-occurring methodological features based on explicit full-text reporting: (i) validation strategy and leakage risk and (ii) multiplicity handling. Validation was categorized as none (apparent in-sample performance only), resampling-based internal validation, split-sample validation, external validation, or unclear; we separately flagged high leakage risk when selection or tuning (e.g., feature selection, cutpoints, hyperparameters) and evaluation were not clearly separated. Multiplicity handling was coded as none acknowledged, family-wise error control, false discovery rate approaches, or a prespecified primary analysis contrasted with exploratory analyses accompanied by interpretive caveats. Two reviewers coded independently and resolved disagreements by consensus or third-reviewer adjudication.

### Simulation experiments

We conducted two simulation experiments (Monte Carlo computational experiments) under prespecified data-generating mechanisms to quantify design-induced distortions under controlled conditions. Across experiments, we separated (i) the data-generating process, (ii) the study design or analysis pipeline being evaluated, and (iii) the estimands and evaluation metrics (discrimination, calibration, error rates, and effect-size bias). Each scenario was replicated across independent draws; results are summarized as Monte Carlo averages with empirical variability across replicates.

### Simulation experiment 1: Enrichment-related circularity and estimand mismatch

Experiment 1 focuses on enrichment-induced estimand shifts and transport of performance to an unselected population. We simulated a population (*n*_pop_ = 10,000) with latent severity *D* ∼ 𝒩(0,1), biomarker B correlated with *D*(*ρ* ∈ {0,0.2} *in the primary analysis*), and a binary outcome *Y* generated from a logistic model depending on *D* with marginal prevalence ≈ 0.30. We compared a global random-sample design (*n* ≤ 2,000) to a biomarker-enriched design that restricted analysis to the top fraction of the biomarker distribution (primary enrichment fraction *f* = 0.20), downsampling to match *n* when necessary. In the grid extension, models fit in enriched samples were additionally evaluated when transported to an independent unselected population draw. We recorded the biomarker-outcome association (Wald *p*-value and slope estimate), discrimination (AUC), overall accuracy (Brier score), and calibration metrics including mean calibration bias and expected calibration error (ECE) computed using 10 quantile bins (ECE10). Under the null (*ρ* = 0), we estimated empirical type I error as *P*(*p* < 0.05).

### Simulation experiment 2: Selection-related circularity (double dipping) and split-sample confirmation

Experiment 2 focuses on selection-inference reuse when screening many candidates and contrasts double dipping with independent confirmation. This simulation experiment quantified false-positive inflation and effect-size exaggeration when many candidate biomarkers are screened and the same dataset is reused for both selection and inference (“double dipping”), compared with an independent confirmation strategy. We generated *p* independent candidate biomarkers *B* ∈ *R*^*n*^ ^×*p*^ with standard normal entries and a continuous outcome *Y* under (i) a global null (*Y* independent of all biomarkers) and (ii) a one-true-biomarker model (*Y* = *β*_true_*B*_1_ + *ε*, *ε* ∼ 𝒩(0,1)). In the double-dip pipeline, we screened all biomarkers in a single dataset using univariable regression/t-tests, selected the smallest p-value, and reported the corresponding p-value and effect estimate as if prespecified. In the split-sample pipeline, selection was performed in a discovery dataset and inference in an independent confirmation dataset generated from the same mechanism. We summarized empirical false-positive rates under the null (*P*(*p* < 0.05) for the reported selected biomarker), selection accuracy in the one-true-biomarker setting, and effect-size inflation (winner’s-curse bias) conditional on selecting the true biomarker. Sensitivity analyses varied the number of candidates *p*, sample size *n*, and true effect size *β*_true_.

## Results

### Corpus and audit flow

The two complementary PubMed searches (trial-focused and trial-readiness-focused) identified 76 unique records after deduplication. Our PubMed queries targeted 2020-2024 records; the final included set contained one online-ahead-of-print report indexed with a 2025 year in PubMed. All 76 were screened, and 76 full texts were sought; 3 reports could not be retrieved. Seventy-three full texts were assessed for eligibility, of which 1 was excluded (non-empirical opinion piece), yielding 72 included studies for the manual audit (**Figure 1**). Among the 72 included studies, 50 (69.4%) were trial-ready cohorts or screening registries, 19 (26.4%) were biomarker-enriched interventional trials, and 3 (4.2%) contributed to both categories (interventional trial results embedded within trial-ready infrastructure). Fluid and imaging biomarkers were widely represented: 46/72 (63.9%) studies reported fluid biomarkers (39/72 [54.2%] plasma/CSF amyloid or tau), while 34/72 (47.2%) incorporated imaging biomarkers (32/72 [44.4%] PET; 30/72 [41.7%] amyloid PET; 13/72 [18.1%] tau PET; 5/72 [6.9%] MRI). Biomarkers were commonly used for eligibility/enrichment (49/72 [68.1%]) and in prognostic/predictive modeling (54/72 [75.0%]), whereas biomarker endpoints were less frequently reported (7/72 [9.7%] primary/secondary; 6/72 [8.3%] primary). Study characteristics and biomarker roles are summarized in **Table 1**.

**Table 1.** Study characteristics and biomarker roles.

| **Domain** | **Characteristic** | **n (%)** |
| --- | --- | --- |
| **Study set** | Included studies | 72 (100.0) |
|  | Trial-ready cohorts/registries only | 50 (69.4) |
|  | Biomarker-enriched interventional trials only | 19 (26.4) |
|  | Both query slices (overlap) | 3 (4.2) |
| **Biomarkers (modalities)** | Any fluid biomarker (plasma and/or CSF) | 46 (63.9) |
|  | Plasma/CSF amyloid or tau biomarkers | 39 (54.2) |
|  | Plasma biomarkers | 39 (54.2) |
|  | CSF biomarkers | 17 (23.6) |
|  | Any imaging biomarker (PET and/or MRI) | 34 (47.2) |
|  | Any PET imaging | 32 (44.4) |
|  | Amyloid PET | 30 (41.7) |
|  | Tau PET | 13 (18.1) |
|  | FDG PET | 3 (4.2) |
|  | MRI | 5 (6.9) |
|  | Both fluid and imaging biomarkers | 25 (34.7) |
| **Biomarkers (targets)** | Any amyloid biomarker (fluid and/or imaging) | 50 (69.4) |
|  | Any tau biomarker (fluid and/or imaging) | 26 (36.1) |
| **Biomarkers (roles)** | Biomarker used for eligibility/enrichment (as extracted) | 49 (68.1) |
|  | Prognostic/predictive modeling mentioned | 54 (75.0) |
|  | Biomarker endpoint mentioned (primary/secondary) | 7 (9.7) |
|  | Biomarker as primary endpoint mentioned | 6 (8.3) |
|  | Clinical/demographic covariates mentioned with biomarkers | 35 (48.6) |

Counts are not mutually exclusive across biomarker modalities, targets, and roles; percentages are calculated with denominator n = 72.

### Prevalence and patterns of circularity

#### Frequency of circular and non-circular designs

On full-text review, 62/72 (86.1%) studies were classified as apparently independent, 7/72 (9.7%) as possibly non-independent, and 3/72 (4.2%) as definitely non-independent. In this corpus, ‘possible’ non-independence most commonly reflected enrichment/range restriction or threshold-based subgrouping (Simulation experiment 1-like) and, less commonly, data-driven selection followed by inference in the same dataset (Simulation experiment 2-like); one ‘possible’ case was rated unclear due to insufficient reporting detail. ‘Definite’ non-independence was assigned when biomarker information was clearly used both to define or strongly enrich the analyzed sample and to support primary inferential claims (for example, biomarker-defined eligibility paired with biomarker-based primary outcomes or prognostic claims). Overall, these findings suggest that circularity is not pervasive, but it is non-trivial in frequency: roughly one in seven recent AD biomarker trials or trial-ready cohorts in the sample raised at least possible concerns about re-using the same biomarker information for both selection and inference.

#### Dominant circularity patterns among flagged studies

Among the 10 studies classified as definitely or possibly non-independent, 7/10 (70%) showed primarily enrichment-related non-independence (Simulation experiment 1-like), where biomarker values or biomarker-derived status were used to define or strongly enrich the analyzed sample and downstream association/performance estimates risked being interpreted beyond that conditional (selected) population. One study (1/10, 10%) showed primarily selection-related non-independence (Simulation experiment 2-like), characterized by data-driven selection (e.g., optimized thresholding or winner-selection) with performance/inference assessed in the same dataset without independent confirmation. One study (1/10, 10%) was classified as hybrid, exhibiting both enrichment-based sample restriction and within-dataset selection. One study (1/10, 10%) was coded as unclear due to insufficient methodological detail to determine the dominant mechanism.

### Co-occurring design and analysis features

We next summarized two methodological features that can amplify circularity-related bias: validation/leakage risk and multiplicity handling.

#### Validation and leakage risk

Among the 72 audited studies, 40/72 (55.6%) reported no explicit validation beyond apparent in-sample performance, and 15/72 (20.8%) described validation procedures too vaguely to classify. Only 17/72 (23.6%) reported an explicit validation strategy: 9/72 (12.5%) used resampling-based internal validation (cross-validation), 4/72 (5.6%) used split-sample validation, and 4/72 (5.6%) reported external validation. Potential information leakage was judged high in 35/72 (48.6%), low in 7/72 (9.7%), and unclear in 30/72 (41.7%), most often because key details (e.g., cutpoint selection, feature selection, or hyperparameter tuning) were not described with sufficient separation between model development and evaluation (**Table 2**).

**Table 2.** Validation, leakage risk, and multiplicity handling.

| Domain | Category | n (%) |
| --- | --- | --- |
| Validation approach (as reported) | No validation (apparent in-sample performance only) | 40 (55.6) |
|  | Unclear / insufficient detail to classify | 15 (20.8) |
|  | Resampling-based internal validation (cross-validation/bootstrapping) | 9 (12.5) |
|  | Split-sample internal validation (train/test split) | 4 (5.6) |
|  | External validation (independent cohort) | 4 (5.6) |
| **Information leakage risk (as judged from reporting)** | High (selection/tuning and evaluation not clearly separated) | 35 (48.6) |
|  | Unclear (insufficient detail to assess) | 30 (41.7) |
|  | Low (clear separation of tuning/selection and evaluation) | 7 (9.7) |
| **Multiplicity handling (multiple comparisons)** | No adjustment and no explicit acknowledgment | 33 (45.8) |
|  | Adjusted for multiple comparisons (FWER or FDR methods) | 20 (27.8) |
|  | Prespecified primary analysis; secondary analyses framed as exploratory | 16 (22.2) |
|  | Unclear / insufficient detail to classify | 3 (4.2) |

Percentages use denominator $n = 72$ ; categories are mutually exclusive within each domain.

#### Multiplicity handling

Multiplicity was frequently a concern: 33/72 (45.8%) explicitly reported multiple inferential endpoints/subgroups without a described adjustment, whereas 20/72 (27.8%) reported a formal adjustment (e.g., family-wise error control or false discovery rate). In 16/72 (22.2%), multiplicity was coded as not present (a single prespecified primary inferential endpoint), and in 3/72 (4.2%) multiplicity handling could not be determined from the full text (**Table 2**).

**Table 3** contextualizes circularity classifications by summarizing, within each circularity stratum, reported validation approaches, judged leakage risk (based on reporting), and multiplicity handling. Even among studies classified as apparently independent, most reported no validation beyond apparent in-sample performance (36/62, 58.1%), whereas split-sample and external validation were uncommon (each 4/62, 6.5%). In the definitely and possibly non-independent strata, validation was more often unclear (2/3, 66.7% and 3/7, 42.9%), reflecting limited methodological detail in a subset of flagged studies. Across strata, leakage risk was rarely judged low: high or unclear leakage risk predominated among apparently independent studies (55/62, 88.7%), and no study in the definitely or possibly non-independent strata was classified as low leakage risk. Multiplicity handling was similarly variable: 29/62 (46.8%) apparently independent studies were coded as unadjusted, while in the flagged strata multiplicity was split across unadjusted, not present, and unclear categories (with small cell counts in the definitely non-independent stratum). Overall, these cross-tabs suggest that limited validation reporting, potential leakage, and incomplete multiplicity handling are common in the corpus and may co-occur even when studies do not meet the prespecified circularity criteria.

**Table 3.** Circularity status vs validation, leakage risk, and multiplicity handling.

(A) Validation strategy
| Circularity status | None | Unclear | Resampling-based internal | Split-sample internal | External |
| --- | --- | --- | --- | --- | --- |
| Independent (n=62) | 36 (58.1) | 10 (16.1) | 8 (12.9) | 4 (6.5) | 4 (6.5) |
| Definite (n=3) | 1 (33.3) | 2 (66.7) | 0 (0.0) | 0 (0.0) | 0 (0.0) |
| Possible (n=7) | 3 (42.9) | 3 (42.9) | 1 (14.3) | 0 (0.0) | 0 (0.0) |

(B) Leakage risk
| Circularity status | High | Unclear | Low |
| --- | --- | --- | --- |
| Independent (n=62) | 30 (48.4) | 25 (40.3) | 7 (11.3) |
| Definite (n=3) | 1 (33.3) | 2 (66.7) | 0 (0.0) |
| Possible (n=7) | 4 (57.1) | 3 (42.9) | 0 (0.0) |

(C) Multiplicity handling
| Circularity status | Unadjusted / not acknowledged | Adjusted (FWER/FDR) | Not present (single prespecified primary analysis) | Unclear |
| --- | --- | --- | --- | --- |
| Independent (n=62) | 29 (46.8) | 19 (30.6) | 12 (19.4) | 2 (3.2) |
| Definite (n=3) | 1 (33.3) | 0 (0.0) | 1 (33.3) | 1 (33.3) |
| Possible (n=7) | 3 (42.9) | 1 (14.3) | 3 (42.9) | 0 (0.0) |

Cells show n (row %). “Independent” = apparently independent/safe; “Possible” = possibly non-independent; “Definite” = definitely non-independent. Percentages are row-wise within circularity strata. “Unclear” indicates insufficient full-text reporting to classify. Counts are descriptive; small cell sizes limit inference.

Across the corpus, circularity concerns and the two co-occurring features rarely appeared in isolation. Studies flagged as definite or possible non-independence more often coincided with limited validation evidence, potential leakage risk, and/or minimal attention to multiplicity; however, these features were also observed in a substantial fraction of studies classified as apparently independent. Taken together, the audit suggests that design-induced non-independence is one identifiable mechanism among broader patterns that can lead to over-optimistic biomarker inferences in contemporary AD biomarker and trial-readiness research.

### Simulation experiment 1: enrichment-related circularity primarily shifts the estimand

Simulation experiment 1 assessed how biomarker-based enrichment changes apparent biomarker performance when the same biomarker both defines the analyzed sample and is used to quantify prognostic performance (**Figures 2 and 3**).

**Figure 2.**
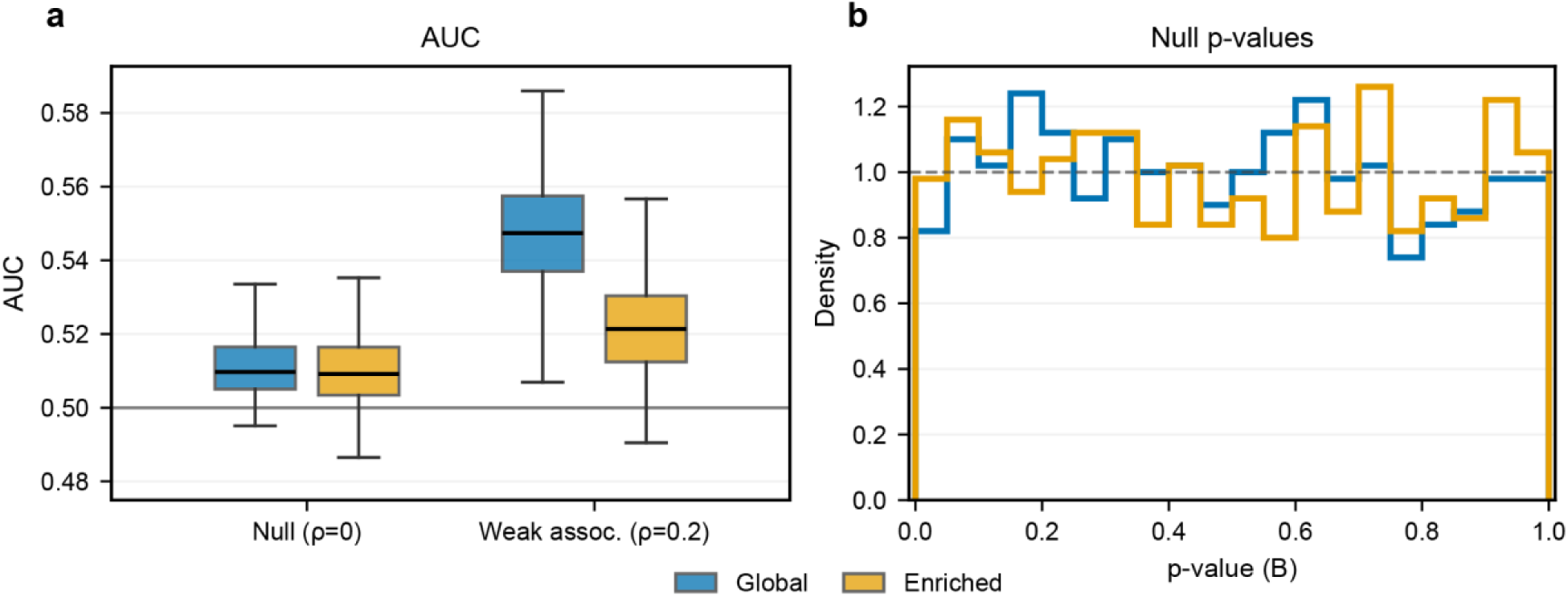
Simulation Experiment 1: Simulation experiment 1: biomarker-based enrichment preserves null error rates but shifts apparent performance under signal. (a) Discrimination (AUC) for biomarker B under two designs: Global (random sample from the full simulated population) and Enriched (restricted to individuals in the top 20% of B; f = 0.20). Results are shown for a null scenario (*ρ* = 0) and a modest-association scenario (*ρ* = 0.2) (1,000 Monte Carlo replicates per scenario). Under the null, mean AUC is similar across designs (≈0.51). Under (*ρ* = 0.2), mean AUC is lower in the enriched design (0.5219) than in the global design (0.5475). (b) Distributions of Wald p-values for the biomarker slope under *ρ* = 0. P-values are approximately uniform in both designs and empirical type I error at *α* = 0.05 is near nominal (0.041 global; 0.049 enriched), indicating that enrichment alone does not induce false-positive inflation.

**Figure 3.**
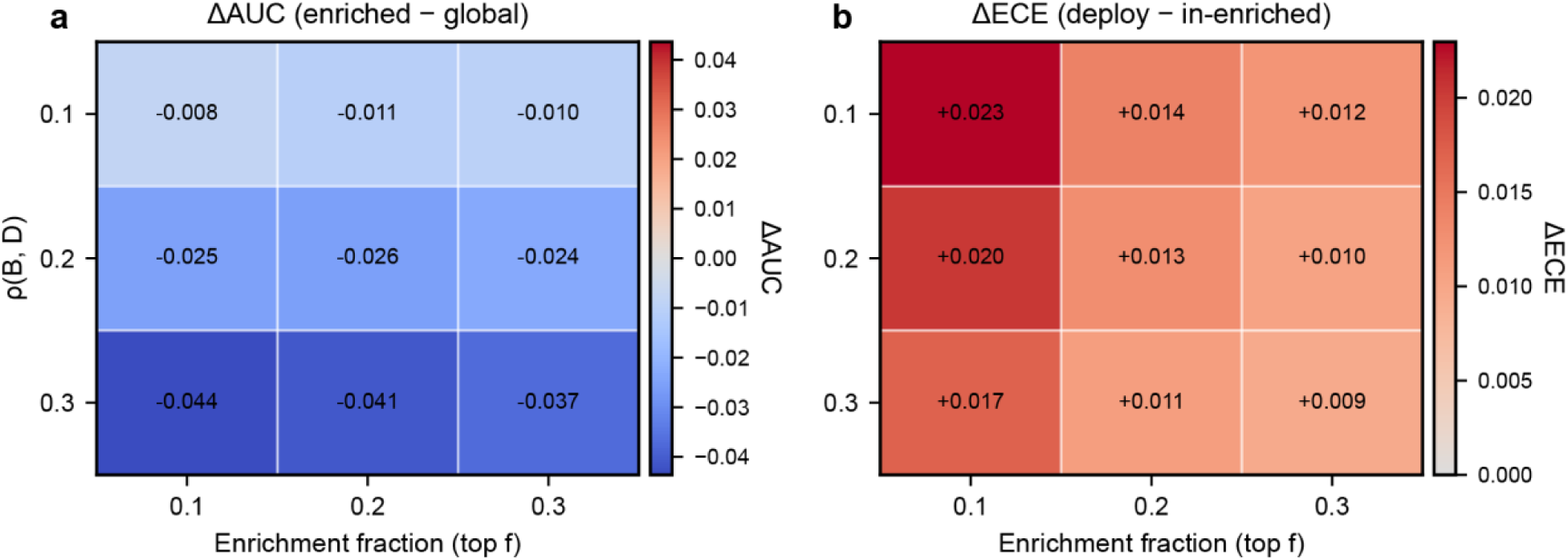
Simulation experiment 1: enrichment attenuates discrimination across scenarios and worsens calibration under transport. (a) Difference in discrimination across a grid of biomarker-disease correlations (*ρ* ∈ {0.1,0.2,0.3})and enrichment fractions (*f* ∈ {0.10,0.20,0.30}). Each cell reports ΔAUC = AUC_enriched_ − AUC_global_ averaged over Monte Carlo replicates; ΔAUC is consistently negative and becomes more negative with stronger association and more stringent enrichment. (b) Calibration drift when models fit in enriched samples are evaluated in an independent unselected population draw. Each cell reports ΔECE = ECE_deploy_ − ECE_enriched_, where ECE is computed using deciles of predicted risk (ECE10). ΔECE is positive across all scenarios, with the largest drift under the most stringent enrichment (*f* = 0.10), consistent with reduced calibration upon transport from enriched trial-like samples to unselected populations.

#### Main scenario (*f* = 0.20): type I error is near-nominal, but performance differs under a true association

Under the null (ρ=0), discrimination was essentially identical in the global and enriched designs (mean AUC 0.511 vs 0.510), and empirical type I error at α=0.05 was close to nominal (0.041 global; 0.049 enriched). In contrast, under a modest association (ρ=0.2), enrichment shifted the estimand: mean AUC decreased from 0.5475 (global) to 0.5219 (enriched), and the biomarker was significant in 92.6% vs 40.2% of simulations, respectively. Calibration error was similar across designs (mean ECE10 0.0234 global vs 0.0239 enriched), but overall prediction error differed, with a lower mean Brier score in the global design (0.2084) than in the enriched design (0.2272) (**Figure 2**).

### Grid of effect sizes and enrichment fractions: discrimination is consistently lower in enriched samples

Across (*ρ* ∈ {0.1,0.2,0.3}) and enrichment fractions (*f* ∈ {0.10,0.20,0.30}), ΔAUC = AUC_enriched_ − AUC_global_ was consistently negative and became more negative with stronger true association. For ρ=0.1, ΔAUC ranged from −0.0080 to −0.0107; for ρ=0.2, from −0.0235 to −0.0258; and for ρ=0.3, from −0.0372 to −0.0436. Differences were generally larger under more aggressive enrichment, though not strictly monotonic in f at 1,000 simulations per grid cell (**Figure 3**).

#### Transport back to an unselected population: calibration degrades with enrichment severity

When models fit in enriched samples were applied to an unselected population, calibration error increased for all (*ρ*, *f*) combinations: ΔECE = ECE_deploy_ − ECE_enriched_ ranged from approximately 0.009 to 0.023, with the largest degradation under the most extreme enrichment (*f* = 0.10) (**Figure 3**).

### Simulation experiment 2: selection-related circularity inflates error rates and effect sizes

Simulation experiment 2 evaluated selection-inference reuse (“double dipping”) when many candidate biomarkers are screened and the strongest association is reported from the same dataset, and compared this to split-sample confirmation (**Figures 4 and 5**).

**Figure 4.**
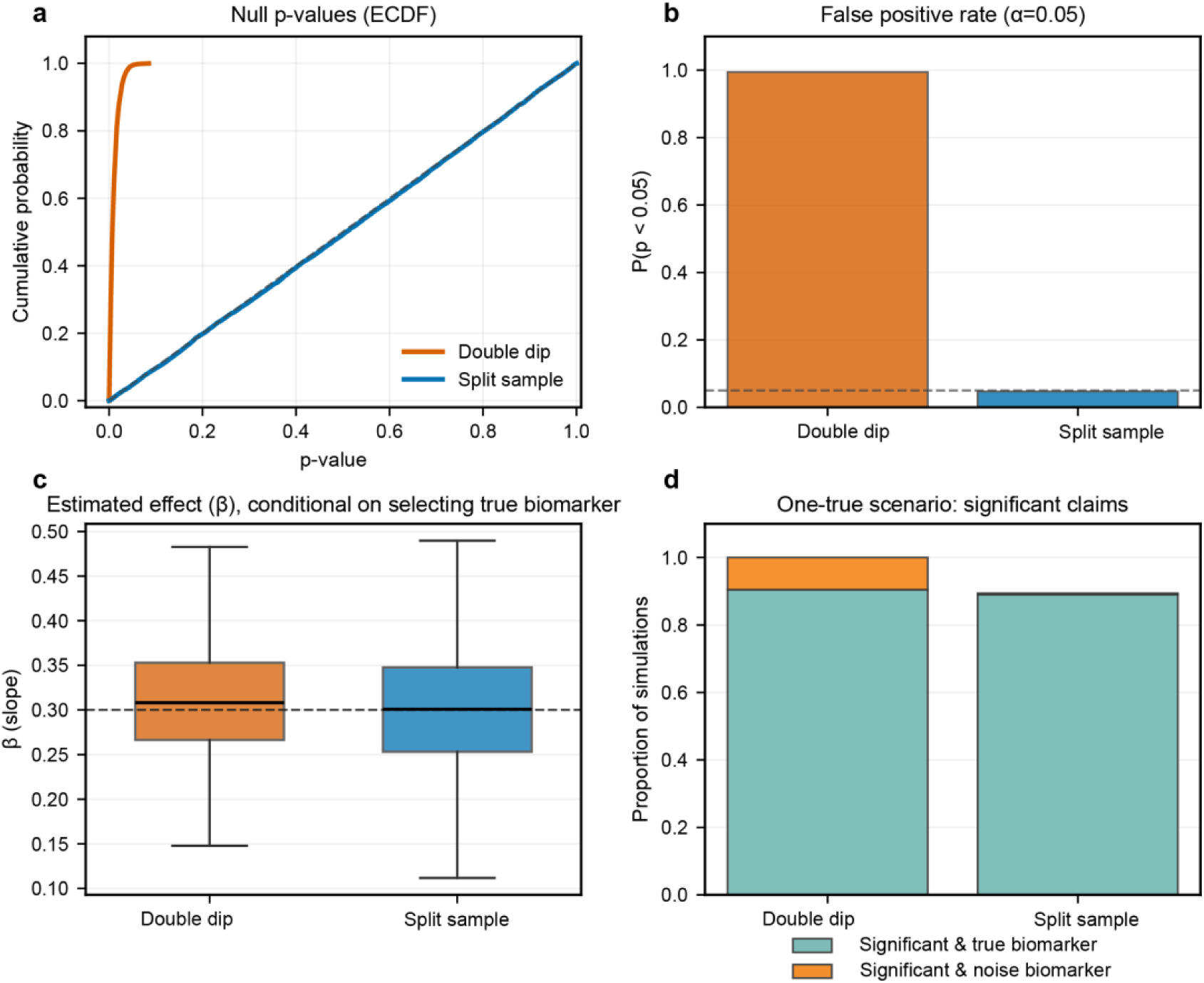
Simulation experiment 2: double dipping yields extreme false-positive inflation and winner’s-curse bias; split-sample confirmation restores nominal behavior. (a) Global null scenario (n=200, p=100; 10,000 Monte Carlo replicates). In the double-dip pipeline (screen p candidates, select the smallest p-value, and report inference from the same dataset), the probability that the selected (“winning”) biomarker met p<0.05 was 0.9943 and winner p-values were concentrated near zero (median 0.00695). In the split-sample pipeline (selection in a discovery set; inference in an independent confirmation set), *P*(*p* < 0.05) = 0.0470 and p-values were approximately uniform (median 0.507). (b) One-true-biomarker scenario (*β*_true_ = 0.3; *n* = 200, *p* = 100). Both pipelines selected the true biomarker at similar rates (0.9045 double-dip; 0.9034 split-sample), but inference differed: the double-dip pipeline produced near-universal significance (overall *P*(*p* < 0.05) = 0.9999) and substantial significance even when a noise biomarker was selected (*P*(*p* < 0.05 ∣ noise selected) = 0.9990), with winner’s-curse inflation in effect estimates when the true biomarker was selected (mean *β̂* = 0.3119). Split-sample confirmation preserved near-nominal behavior when a noise biomarker was selected (*P*(*p* < 0.05 ∣ noise selected) = 0.0414) and yielded essentially unbiased estimates when the true biomarker was selected (mean *β̂* = 0.3003).

**Figure 5.**
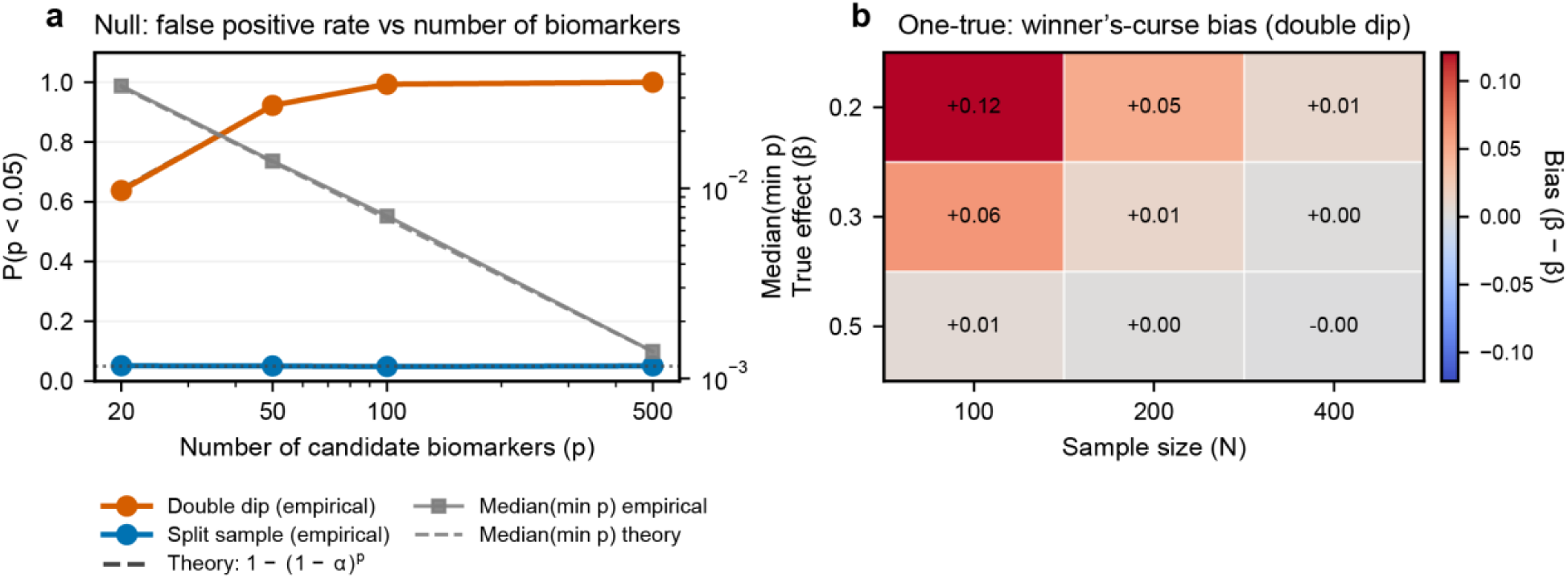
Simulation experiment 2: error inflation scales with the number of candidates and winner’s-curse bias is largest in low-information regimes. (a) Global null scenario (*n* = 200): empirical false-positive rate for the selected biomarker as a function of the number of screened candidates (*p* ∈ {20, 50, 100, 500}). In the double-dip pipeline, the probability that the selected biomarker met *p* < 0.05 increased with p and closely tracked the theoretical family-wise error rate under independence, 1 − (1 − 0.05)^*p*^ (*e*. *g*., 0.6375 *at p* = 20 *and* 1.0000 *at p* = 500). In the split-sample pipeline, false-positive rates remained near 0.05 across all p. (b) One-true-biomarker grid (*n* ∈ {100, 200, 400}, *β*_true_ ∈ {0.2, 0.3, 0.5})conditional effect-size bias for *β̂* given that the true biomarker was selected. Double-dip conditional bias was largest in low-information settings (e.g., *n* = 100, *β*_true_ = 0.2: conditional bias ≈+0.121) and diminished with larger n and stronger signal; split-sample conditional bias remained near zero across the grid.

#### Main scenario (n=200, p=100): near-certain false positives under the null

Under the global null, the probability that the selected (“winning”) biomarker met *p* < 0.05 was 0.9943 in the double-dip pipeline versus 0.0470 with split-sample confirmation; winner p-values were highly concentrated near zero (median 0.00695 vs 0.507). Thus, when many candidates are screened and the most extreme association is re-tested in the same data, false positives become effectively guaranteed at these dimensions (**Figure 4**).

#### One-true-biomarker scenario (*β*_true_ = 0.3): similar selection accuracy, divergent inference

When one true biomarker effect was present, both pipelines selected the true biomarker at similar rates (0.9045 double-dip; 0.9034 split-sample). However, inference diverged. In the double-dip pipeline, the reported association was almost always significant (overall *P*(*p* < 0.05) ≈ 0.9999), including when a noise biomarker was selected (overall *P*(*p* < 0.05) ≈ 0.9999). Conditional on selecting the true biomarker, effect estimates showed winner’s-curse inflation (mean *β̂* ≈ 0.312 *vs β*_true_ = 0.3). In contrast, split-sample confirmation preserved near-nominal behavior when a noise biomarker was selected (*P*(*p* < 0.05 ∣ noise selected) = 0.0414) and yielded essentially unbiased estimates when the true biomarker was selected (mean *β̂* ≈ 0.300) (**Figure 4**).

#### Sensitivity analyses: inflation scales with p; split-sample is stable

Under the null, double-dip false-positive rates tracked the expected family-wise error rate 1 − (1 − 0.05)^*p*^, rising from 0.6375 at *p* = 20 to ≈ 1.0000 at *p* = 500, whereas split-sample confirmation remained near 0.05 across all p (**Figure 5**). In a grid of one-true-biomarker scenarios (*n* ∈ {100,200,400}, *β*_true_ ∈ {0.2,0.3,0.5}), winner’s-curse bias in the double-dip pipeline was largest in low-information settings (e.g., *n* = 100, *β*_true_ = 0.2: conditional bias ≈+0.121) and diminished with increasing sample size and signal strength; split-sample conditional bias remained near zero across the grid (**Figure 5**).

### Synthesis across audit and simulation experiments

Across the audited corpus, non-independence, when present, most commonly reflected enrichment-related designs, in which biomarker-defined inclusion or subgrouping was followed by association or performance claims that could be misinterpreted as applying to a broader, unselected clinical population. Consistent with this empirical pattern, Simulation experiment 1 showed that enrichment did not materially inflate type I error under the null, but it shifted the estimand: discrimination was consistently lower in enriched samples under a true association, and calibration degraded when models fit under enrichment were deployed to an unselected population. In contrast, selection-related non-independence was less frequent in the audit but, as demonstrated by Simulation experiment 2, produced extreme false-positive inflation and winner’s-curse bias when many candidates were screened and inference was performed on the same dataset. Together, these results delineate two distinct pathways by which biomarker-centered study designs can yield over-optimistic inferences, motivating the reporting and design recommendations developed below.

## Discussion

As AD biomarkers transition from primarily research measures to determinants of trial eligibility and clinical decision-making (via biomarker-confirmed treatment pathways), design choices that condition on biomarker status increasingly shape who is studied and how findings generalize [6, 32–34]. In pivotal anti-amyloid trials, biomarker confirmation is used to define trial populations, underscoring that biomarker-based inclusion is now a routine and consequential design feature [7, 35–37]. Against this backdrop, we asked a methodological question with direct interpretive stakes: when biomarkers are used to define (or strongly enrich) the analyzed sample, or when many candidate biomarkers/regions are screened and the ‘best’ signal is re-tested in the same dataset, how do these choices bias reported prognostic or surrogate claims [16, 19, 22, 37]. Our audit and simulations delineate two avoidable mechanisms, enrichment-driven estimand mismatch and selection-inference reuse (‘double dipping’), that can materially distort inference unless discovery/confirmation roles, validation, and multiplicity control are made explicit [16, 19, 22].

In our full-text audit of 72 recent AD biomarker trials and trial-ready cohorts, most studies were classified as apparently independent with respect to prespecified circularity criteria (62/72, 86.1%), but a non-trivial minority raised definite or possible concerns about non-independence (10/72, 13.9%; **Table 1 and 3**).

Among flagged studies, the dominant mechanism was enrichment-related non-independence (7/10), where biomarker-defined inclusion, thresholding, or subgrouping restricts the analysis set in ways that can shift the estimand and invite over-generalization if results are interpreted as reflecting performance in the broader clinic population (see Results section). Selection-inference reuse (‘double dipping’) was less frequent in absolute terms (1/10 primary; 1/10 hybrid), but its presence is notable because screening many candidate biomarkers or regions and then reporting inference from the same dataset is a well-established pathway to inflated evidence if not paired with independent confirmation or appropriate multiplicity control [16, 38].

Importantly, the circularity patterns we flagged rarely occurred in isolation. Across the full corpus, key safeguards that determine how interpretable and how transportable biomarker claims are (out-of-sample validation, clear separation of model selection/tuning from evaluation to limit information leakage, and explicit multiplicity handling) were frequently absent or insufficiently reported (**Table 3**). More than half of studies reported no validation beyond apparent in-sample performance (40/72, 55.6%), and leakage risk was judged high or could not be assessed due to incomplete reporting in 65/72 (90.3%). These vulnerabilities were also common among studies we classified as apparently independent (e.g., 36/62 [58.1%] with no reported validation; 55/62 [88.7%] with high or unclear leakage risk; 29/62 [46.8%] with no stated multiplicity adjustment), supporting the view that non-independence is one identifiable mechanism within a broader set of routine design-and-reporting choices that can undermine reproducibility and transportability. Because our labels reflect what was explicitly stated in full text, the substantial “unclear” fractions should be interpreted primarily as limited transparency rather than evidence of flawed conduct; nonetheless, inadequate reporting prevents readers from distinguishing robust out-of-sample evidence from potentially optimistic, selection-contaminated estimates, concerns explicitly emphasized in prediction-model reporting and appraisal guidance (TRIPOD [22]; PROBAST [26]), and in methodological work showing that performance estimates are biased when tuning/feature selection is not properly nested within validation or when the same data are reused for selection and inference (“double dipping”) [16–18]. Finally, where many biomarkers, regions, or derived scores are tested, unaddressed multiplicity predictably increases false-positive risk; accepted controls include family-wise error procedures and false discovery rate approaches [25, 39].

When biomarkers define eligibility or analyses are restricted post hoc to “biomarker-high” subsets then the resulting association and performance estimates are conditional on a deliberately altered case mix and therefore target a different estimand than the unselected clinic population from which participants are drawn [16, 37, 38]. This distinction matters because both discrimination and calibration are known to vary with case mix, baseline risk, and predictor distributions; performance assessed in a restricted or enriched sample may not transport unchanged to the broader population without additional evidence or adjustment [20]. Simulation experiment 1 isolates this mechanism: biomarker-based enrichment does not, by itself, induce false-positive inflation under the null, but under a modest true association it systematically changes apparent discrimination within the enriched subset and yields calibration drift when an enriched-fit model is deployed to an independent, unselected population (**Figures 2, 3**). Accordingly, biomarker-enriched trials and trial-ready infrastructures can provide valid inferences for the enriched population they are designed to study, but manuscripts should (i) state the target population/estimand explicitly and (ii) reserve broader prognostic or surrogate generalizations for settings supported by external validation or principled transport/model-updating (recalibration) analyses [22, 26, 28, 29].

Selection-related circularity is qualitatively different: it arises when the analysis lets the dataset determine which biomarker/region/panel is “most associated” and then reports conventional p-values and effect estimates for that selected feature as if the hypothesis had been prespecified. Because selection and inference are no longer independent, nominal p-values no longer control type I error, and false-positive rates increase rapidly with the number of candidates screened [16, 17, 19, 39]. Simulation experiment 2 isolates this mechanism: under a global null with *n* = 200 and *p* = 100, the double-dip pipeline yielded near-certain “significance” for the selected biomarker (*P*(*p* < 0.05) = 0.994), closely matching the theoretical family-wise error 1 − (1 − 0.05)^*p*^, whereas split-sample confirmation remained near nominal (0.047) with approximately uniform p-values (median 0.507). Under a one-true-biomarker setting (*β*_true_ = 0.3), both pipelines selected the true biomarker at similar rates (∼0.90), but same-sample inference converted both true and spurious “wins” into near-universal statistical significance and produced effect-size inflation (winner’s curse); split-sample confirmation substantially mitigated both by re-estimating the selected association in independent data (**Figures 4, 5**).

A constructive interpretation of these findings is that the main vulnerabilities we observed are, in principle, addressable through clearer alignment between study aims, estimands, and reporting, rather than through novel statistical machinery [40, 41]. In particular, biomarker-enriched AD trials and trial-ready infrastructures often combine elements of randomized trials (eligibility/enrichment and endpoints), diagnostic evaluation (thresholds and classification), and prognostic modeling (risk prediction and performance reporting). Established reporting and appraisal frameworks already provide concrete expectations for these components, e.g., transparent specification of populations, outcomes, modeling steps, validation, and multiplicity, but they are rarely applied explicitly as a unified standard in biomarker-enriched studies [22, 26, 40, 41]. Positioning enriched-sample results as conditional (rather than implicitly population-wide), separating discovery from confirmation when screening many candidates, and reporting validation and leakage safeguards in sufficient detail would allow readers to distinguish (i) valid within-design inferences from (ii) claims that require additional transport or independent confirmation. This perspective is deliberately collaboration-forward: biomarker enrichment and high-dimensional exploration are often necessary and have proven to be scientifically valuable in AD research, but they warrant correspondingly careful language about generalizability and clear evidence of out-of-sample evaluation when claims are intended to inform broader clinical or regulatory interpretation.

Translating these observations into practice does not require new methodology so much as tighter adherence to established guidance. First, manuscripts should state the target population and estimand explicitly, i.e., whether reported associations and performance are intended to apply within a biomarker-enriched trial population or to the broader clinic-source population from which that trial population was drawn (and therefore require transport/validation) [14, 22, 26]. Second, when many candidate biomarkers or regions are screened, exploratory selection should be separated from confirmatory inference (e.g., split-sample or external confirmation), and any performance estimation should nest all selection/tuning steps to avoid optimistic bias from selection-inference reuse (“double dipping”) and leakage [16–18]. Third, for any prognostic or surrogate-performance claim, authors should report (operationally, not aspirationally) the validation strategy and multiplicity plan, consistent with prediction-model reporting and risk-of-bias frameworks, because in contemporary AD trials, amyloid and/or tau confirmation is often an explicit eligibility requirement, making it easy for readers to over-transport enriched-sample results unless conditional vs transportable claims are clearly delimited [7, 25, 26, 36, 39].

### Limitations

Several limitations bound interpretation of both the audit and the simulation experiments. First, the audited corpus was defined by two prespecified, targeted PubMed (MEDLINE) queries (2020-2024) and was restricted to records with accessible full text; accordingly, the resulting frequencies should be interpreted as descriptive of this sampling frame rather than an exhaustive census of AD biomarker studies across all venues and indexing services. Second, our coding reflects what was explicitly reported in the article full text: “unclear” categories primarily index limits of reporting transparency (what an external reader can verify), not a determination that a safeguard was absent; our inferences therefore emphasize interpretability and transportability of the reported evidence rather than intent or conduct.

Third, the simulation experiments were deliberately stylized to isolate two generic mechanisms, range restriction/enrichment-induced estimand shift and selection-inference reuse (“double dipping”) and were not intended to reproduce the full complexity of AD endpoints, correlated multimodal biomarkers, longitudinal designs, or adaptive recruitment. This scope is consistent with established guidance that simulation studies function as controlled “*in-sillico* experiments” for clarifying statistical behavior under prespecified data-generating mechanisms, prioritizing mechanistic insight over disease-specific parameter calibration [42, 43].

### Conclusions

In summary, our audit and simulation experiments delineate two common, and largely avoidable, routes to over-optimistic biomarker inference in contemporary AD trial and trial-readiness research. First, biomarker-based enrichment can induce an estimand mismatch, in which performance estimated conditional on biomarker-defined eligibility is later discussed as if it were the biomarker’s performance in the broader, clinic-source population; this is fundamentally an estimand/target-population problem and is best addressed by explicit specification of the estimand and its target population (and, where feasible, separate reporting for the enriched versus source population) [14]. Second, selection-inference reuse (“double dipping”), screening many candidate markers and then reporting inference on the “winner” from the same dataset, can invalidate nominal p-values and inflate effect sizes, a phenomenon extensively described across high-dimensional biomedical and neuroimaging settings [16–18]. The methodological opportunity is therefore not to abandon biomarker-enriched or high-dimensional exploratory designs, but to make their inferential scope explicit and reproducible through simple safeguards: (i) pre-specify and report the estimand and target population, (ii) separate discovery from confirmation (via split-sample or external confirmation), and (iii) ensure that all selection/tuning occurs within validation loops (e.g., nested resampling) or is quarantined from final evaluation; when multiple candidates are tested, incorporate an explicit multiplicity strategy aligned to the study’s inferential intent [22, 25, 26, 41].

## Author contributions

S.K. and K.V. conceived and designed the study. S.K. developed the simulation framework, implemented and executed the simulation experiments, and generated figures and quantitative summaries. S.K. and K.V. independently performed full-text review and coding using prespecified criteria, resolved discrepancies by consensus, and curated the extracted variables. S.K. performed the statistical analyses and drafted the manuscript. S.K. and K.V. contributed to interpretation of findings and critically revised the manuscript for intellectual content. Both authors approved the final version of the manuscript and agree to be accountable for all aspects of the work.

## Statements and declarations

### Ethical considerations

This article does not contain any studies with human or animal participants. There are no human participants in this article and informed consent is not required.

### Funding Statement

The author(s) received no financial support for the research, authorship, and/or publication of this article.

### Data availability

All PubMed search queries used to define the audit sampling frame, together with the full simulation code and figure-generation scripts needed to reproduce the experiments, are available at https://github.com/SatwantKumar/ad-biomarker-circularity-simulations.

### Declaration of conflicting interest

The author(s) declared no potential conflicts of interest with respect to the research, authorship, and/or publication of this article

## Notes

### Competing Interest Statement

The authors have declared no competing interest.

